# Nocturnal Respiratory Rate Dynamics Enable Early Recognition of Impending Hospitalizations

**DOI:** 10.1101/2022.03.10.22272238

**Authors:** Nicholas Harrington, David Torres Barba, Quan M. Bui, Andrew Wassell, Sukhdeep Khurana, Rodrigo B. Rubarth, Kevin Sung, Robert L. Owens, Parag Agnihotri, Kevin R. King

## Abstract

The days and weeks preceding hospitalization are poorly understood because they transpire before patients are seen in conventional clinical care settings. Home health sensors offer opportunities to learn signatures of impending hospitalizations and facilitate early interventions, however the relevant biomarkers are unknown. Nocturnal respiratory rate (NRR) is an activity-independent biomarker that can be measured by adherence-independent sensors in the home bed. Here, we report automated longitudinal monitoring of NRR dynamics in a cohort of high-risk recently hospitalized patients using non-contact mechanical sensors under patients’ home beds. Since the distribution of nocturnal respiratory rates in populations is not well defined, we first quantified it in 2,000 overnight sleep studies from the NHLBI Sleep Heart Health Study. This revealed that interpatient variability was significantly greater than intrapatient variability (NRR variances of 11.7 brpm^2^ and 5.2 brpm^2^ respectively, n=1,844,110 epochs), which motivated the use of patient-specific references when monitoring longitudinally. We then performed adherence-independent longitudinal monitoring in the home beds of 34 high-risk patients and collected raw waveforms (sampled at 80 Hz) and derived quantitative NRR statistics and dynamics across 3,403 patient-nights (n= 4,326,167 epochs). We observed 23 hospitalizations for diverse causes (a 30-day hospitalization rate of 20%). Hospitalized patients had significantly greater NRR deviations from baseline compared to those who were not hospitalized (NRR variances of 3.78 brpm^2^ and 0.84 brpm^2^ respectively, n= 2,920 nights). These deviations were concentrated prior to the clinical event, suggesting that NRR can identify impending hospitalizations. We analyzed alarm threshold tradeoffs and demonstrated that nominal values would detect 11 of the 23 clinical events while only alarming 2 times in non-hospitalized patients. Taken together, our data demonstrate that NRR dynamics change days to weeks in advance of hospitalizations, with longer prodromes associating with volume overload and heart failure, and shorter prodromes associating with acute infections (pneumonia, septic shock, and covid-19), inflammation (diverticulitis), and GI bleeding. In summary, adherence-independent longitudinal NRR monitoring has potential to facilitate early recognition and management of pre-symptomatic disease.

## INTRODUCTION

Hospitalizations are extraordinarily common, costly, and occur for diverse reasons, particularly in those with multiple chronic medical conditions (e.g., heart failure, arrhythmias, recurrent pneumonias, sepsis, COPD). Hospitalizations are associated with increased functional disability and decreased quality of life, particularly when they are repeated and prolonged. Fortunately, many hospitalizations are avoidable with early recognition and intervention *(1-10)*.

In the outpatient setting, providers are not aware of clinical deterioration until the patient, family, or caregiver recognizes and reports symptoms. Often, the first recognition by a provider occurs when the patient presents to the emergency room. Symptoms are difficult to self-recognize because they are subjective, and even then are typically lagging indicators of illness *(1)*. Patients are also often reluctant to seek immediate medical attention out of fear they may require hospitalization, a factor that has been aggravated by the recent SARS-CoV-2 pandemic *(2)*. If pre-symptomatic prodromes of hospitalization could be recognized in the home, changes of management or treatment could begin earlier, in lower acuity care settings, potentially reducing the number and duration of hospitalizations and increasing hospital-free-days. The potential for early intervention is particularly attractive for cardiopulmonary diseases such as heart failure, COPD, and pneumonia, which represent the most common causes of hospitalizations in older adults. These patients often present with respiratory symptomatology, and are associated with well-established interventions that can be administered in clinic or at home *(3-6)*. For example, diuretics are often uptitrated for heart failure exacerbations; bronchodilators, steroids, and antimicrobials are administered for COPD exacerbations; and antibiotics are initiated for early uncomplicated bacterial respiratory infections *(4, 5, 7)*. While early recognition of clinical deterioration may be useful, the biomarkers of impending hospitalization are unknown.

Respiratory rate (RR) is a continuously evolving vital sign that is often overlooked, except at its extremes. In awake individuals, its interpretation is complicated by its dependence on voluntary effort, activity level, effort, and emotion *(8, 9)*. However, during sleep nocturnal respiratory rate (NRR) reflects underlying physiologic and pathophysiologic determinants *(10, 11)*. We hypothesized that NRR may be useful as a biomarker of impending hospitalization in the home because respiratory rate strongly impacts inpatient deterioration scores *(12-17)* and because it is readily measured longitudinally by non-contact adherence-independent sensors in the home.

Here, we explore the patterns of NRR and its utility as a biomarker of impending hospitalizations that can be measured longitudinally without requiring patient adherence. We first analyzed NRR distributions in a large cohort of publicly available sleep studies to define the distribution of NRR in populations. Next, we deployed fully automated home bed respiratory monitoring technology to patient homes and defined longitudinal NRR trends without requiring any patient participation. Finally, we examined whether excursions of NRR from patient-specific baselines could be used to detect impending hospitalizations. In a high-risk cohort of 34 patients, we collected and analyzed 4,326,167 epochs at 30-second intervals across 3,403 patient-nights and found that at nominal alarm thresholds, we detected 11 of 23 hospitalizations, with warning windows ranging up to weeks in advance, while only alarming on 2 clinically stable patients who did not need hospitalization. Taken together, this suggests that NRR may be a useful biomarker for automated early detection of impending hospitalizations.

## RESULTS

### Nocturnal respiratory rates in 2,000 single-night sleep studies

Since NRR dynamics are not routinely measured, their distribution in populations or in individuals measured across time is not well characterized. To define NRR in populations, we collected and analyzed raw waveforms from chest belt respirometers of 2,000 polysomnograms from the Sleep Heart Health Study (SHHS), a multi-center cohort study designed to study the cardiovascular and other consequences of sleep-disordered breathing*(18, 19)*. We performed peak-finding and derived time-averaged respiratory rate estimates at 30-second intervals (epochs) as previously described*(20)*. To enable visualization of the entire dataset at-a-glance, we encoded the NRR as color, which enabled the entire sleep study of a patient to be communicated as a sequence of colors assembled as a column read from top to bottom (**Fig. 1a**). To aid visualization, we sorted patients by their average NRR and artificially registered them to a common start time. We quantified 1.84 million epochs across the 2,000-patient dataset and revealed a population mean of 16.4 breaths per minute (brpm), a variance of 11.7 brpm^2^, and range of 31.4 brpm (**Fig. 1b**). By inspection of the heatmap, interpatient variations were subjectively large compared intrapatient variation across time. Based on this observation, we designed the colormap to minimize subjective spectral variation within nights while maximizing variation between patients. To quantitatively compare interpatient and intrapatient variability, we corrected each epoch with a universal NRR mean or patient-specific NRR mean and found that the use of patient-specific references reduced variance by 56% (11.7 brpm^2^ and 5.2 brpm^2^ respectively). This confirmed that interpatient NRR variability was significantly greater than intrapatient NRR variability within a single night (**Fig. 1c**). However, while these data define NRR dynamics during isolated nights (1 per patient), the longitudinal trends of NRR are unknown.

**Figure 1.**
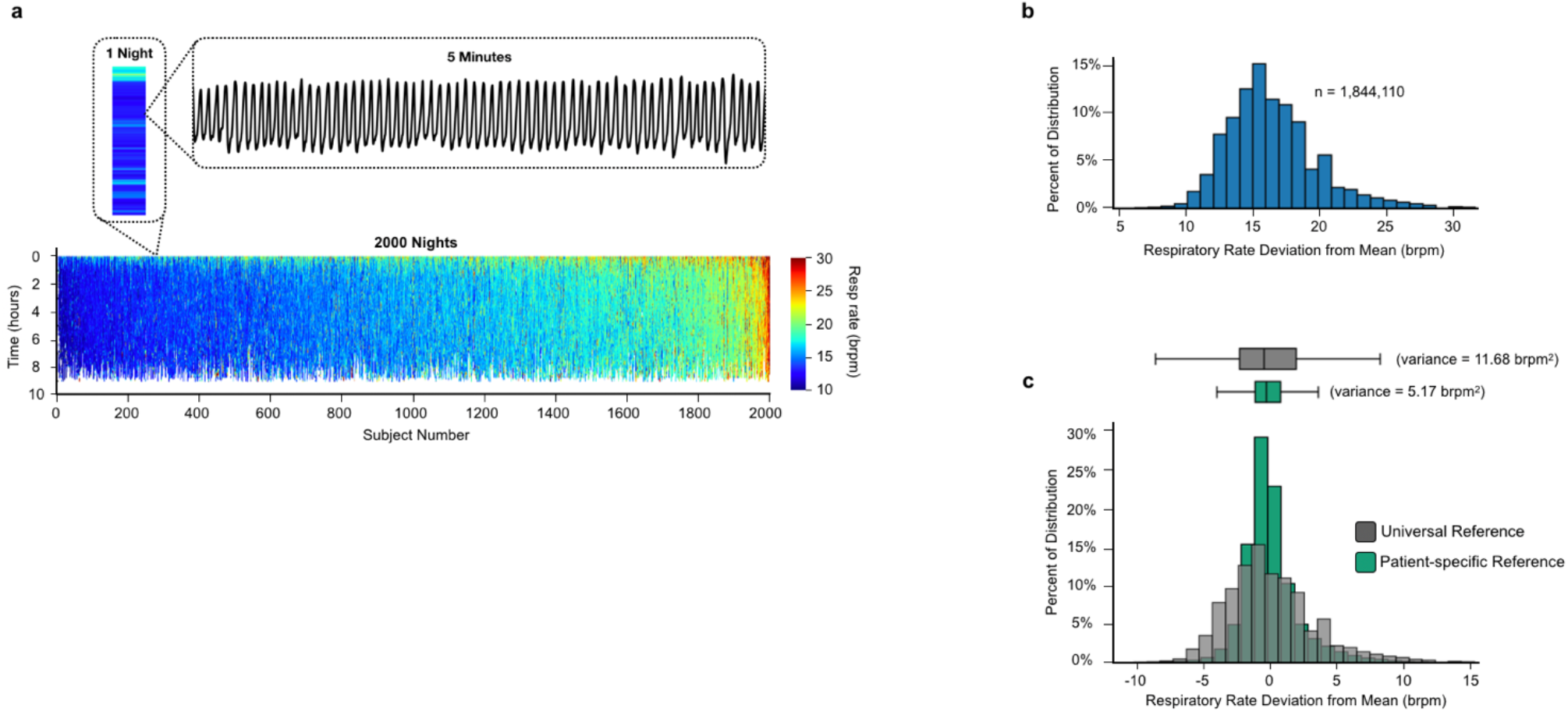
Nocturnal respiratory rate in sleep studies. **a**, Heatmap of 2000 subjects from NHLBI Sleep Heart Health Study. Color indicates respiratory rate. Y-axis is in hours of sleep. X-axis is in subject number. First inset shows 1 night of respiratory rates as single column heatmap. Second inset shows 5 minutes of respiratory waveform. Colorbar ranges from 10-30 brpm, values below 10 brpm not shown. **b**, Histogram of all epochs (n = 1,844,110) from 2000 subjects (mean = 16.36 brpm, variance = 11.68 brpm^2^, range = 31.44 brpm). **c**, Histograms of all epochs centered at the universal mean (gray) and each subject centered on their own mean (green) (variance = 5.17 bprm^2^). The center line in the boxes above the histograms represents the median value, the boxes extend from the first to the third quartiles, and the whiskers extend 1.5x the interquartile range before the first quartile and past the third quartile.

### Adherence-independent monitoring of nocturnal respiratory rates in the home bed

To determine the longitudinal patterns of NRR, we analyzed data from an observational study of adherence-independent home bed monitoring. We selected patients enrolled from 6/22/2020 to 1/20/2021 from an academic health center, cardiology practice, and population health program who were felt by their providers to be at high risk of hospitalization for cardiopulmonary causes in the next year (**Table 1**). Patients signed consent forms and were on average 75 years old and had multiple co-morbidities as detailed in Table 1. This yielded 34 patients and 3403 total patient-nights of monitoring. After removing nights (10%) due to absent or inadequate data, commonly because the patient was away from home or in the hospital, 3,056 patient-days remained for analysis. Participants spanned diverse ages (20’s-90’s), genders, ethnicities, socioeconomic contexts, body weights, and medical comorbidities; settings ranged from transitional housing, studio apartments, and condos, to multi-story homes in gated subdivisions; and sleep furniture included couches, recliners, and a diversity of beds (twins, queens, kings, and adjacent twins) (9 of our patients shared a bed with a partner, see methods for details). Many patients suffered from physical or cognitive disabilities that would have made usage of wearables and mobile apps challenging. For example, we monitored patients with dementia, a woman with blindness from diabetic retinopathy, and another who was bed-bound and neurologically impaired after a debilitating stroke.

**Table 1.** Summary of longitudinal home monitoring patient cohorts.

|  | **Full Cohort**<br>**(34** | **Admission**<br>**(16 patients)** | **No Admission**<br>**(18 patients)** |
| --- | --- | --- | --- |
| **Age (years)** | 75.6 | 74.1 | 76.9 |
| **Gender (Male)** | 23 (67.6%) | 13 (81.3%) | 10 (55.6%) |
| **Race (White)** | 22 (64.7%) | 9 (56.3%) | 13 (72.2%) |
| **CHF** | 27 (79.4%) | 16 (100%) | 11 (61.1%) |
| **LVEF (%)** | 53.1 | 47.8 | 57.8 |
| **Body Mass Index (kg/m2)** | 27.7 | 28.4 | 27.1 |
| **Atrial Fib/flutter** | 22 (64.7%) | 12 (75%) | 10 (55.6%) |
| **Diabetes** | 15 (44.1%) | 7 (43.8%) | 8 (44.4%) |
| **Hypertension** | 29 (85.3%) | 15 (93.8%) | 14 (77.8%) |
| **Hyperlipidemia** | 26 (76.5%) | 13 (81.3%) | 13 (72.2%) |
| **CAD / MI** | 19 (55.9%) | 9 (56.3%) | 10 (55.6%) |
| **COPD** | 12 (35.3%) | 6 (37.5%) | 6 (33.3%) |
| **Stroke/TIA** | 8 (23.5%) | 5 (31.3%) | 3 (16.7%) |
| **Immunosuppressant** | 6 (17.6%) | 4 (25.0%) | 2 (11.1%) |
| **Transplant** | 3 (8.8%) | 2 (12.5%) | 1 (5.6%) |

The sensors were installed under the home beds, plugged into wall power to facilitate indefinite monitoring, and connected to WiFi via custom installation app. Once installed, sensors automatically collected and transmitted data to a cloud environment. No active patient participation was required. Adherence-independent mechanical force measurements were continuously and automatically collected at 80Hz and transmitted hourly to a cloud environment. Respiratory waveforms were quantified using a validated pipeline similar to that used for the sleep studies above*(20)*. NRRs were deposited in a database that was accessible via a password and two-factor authentication-protected web or mobile app. The resulting data set spanned over 3,000 patient-nights and consisted of 4 million NRR epochs. To enable rapid review of longitudinal bed sensor data for each patient, we constructed heatmaps that allowed months of respiratory rates to be rapidly reviewed and interpreted by inspection, “at a glance”, with *Time of day* shown on the y axis (24 hours, from noon-to-noon, divided into 30-second epochs), and successive *Nights* on the x axis. As before, NRRs were displayed as color-coded horizontal bars defined at each 30-second epoch. This allowed each night to be interpreted in the context of months of monitoring.

### Longitudinal NRR in patients with clinical stability

We first divided patients into 2 groups, those who were hospitalized during the period of monitoring and those who were not. We focused first on the patients who were not hospitalized during the analysis period. Like the sleep studies, individual patients were qualitatively similar to themselves across time but different from other patients as illustrated by four examples (**Fig.2a-d**). To analyze the characteristics of stable patients, and to do so in a manner that equally weighted each patient regardless of monitoring duration, we chose 30 nights from each stable patient generating a distribution of over 600,000 epochs, with a mean of 17 brpm and a variance of 16 brpm^2^ (**Fig.2e**). Quantitatively, when the distribution of NRRs for individuals were corrected using a universal reference, the variance was significantly greater (15.9 brpm^2^) than when corrected using a patient-specific reference (7.2 brpm^2^) (**Fig.2f**). This suggested that longitudinal variation for individual patients is low in clinically stable patients. It also suggested that from a practical perspective, detection of excursions from normal should compare to patient-specific NRR baselines rather than universal population norms, as the interpatient variation is in general larger than the intrapatient variation **(Fig. 2g)**.

**Figure 2.**
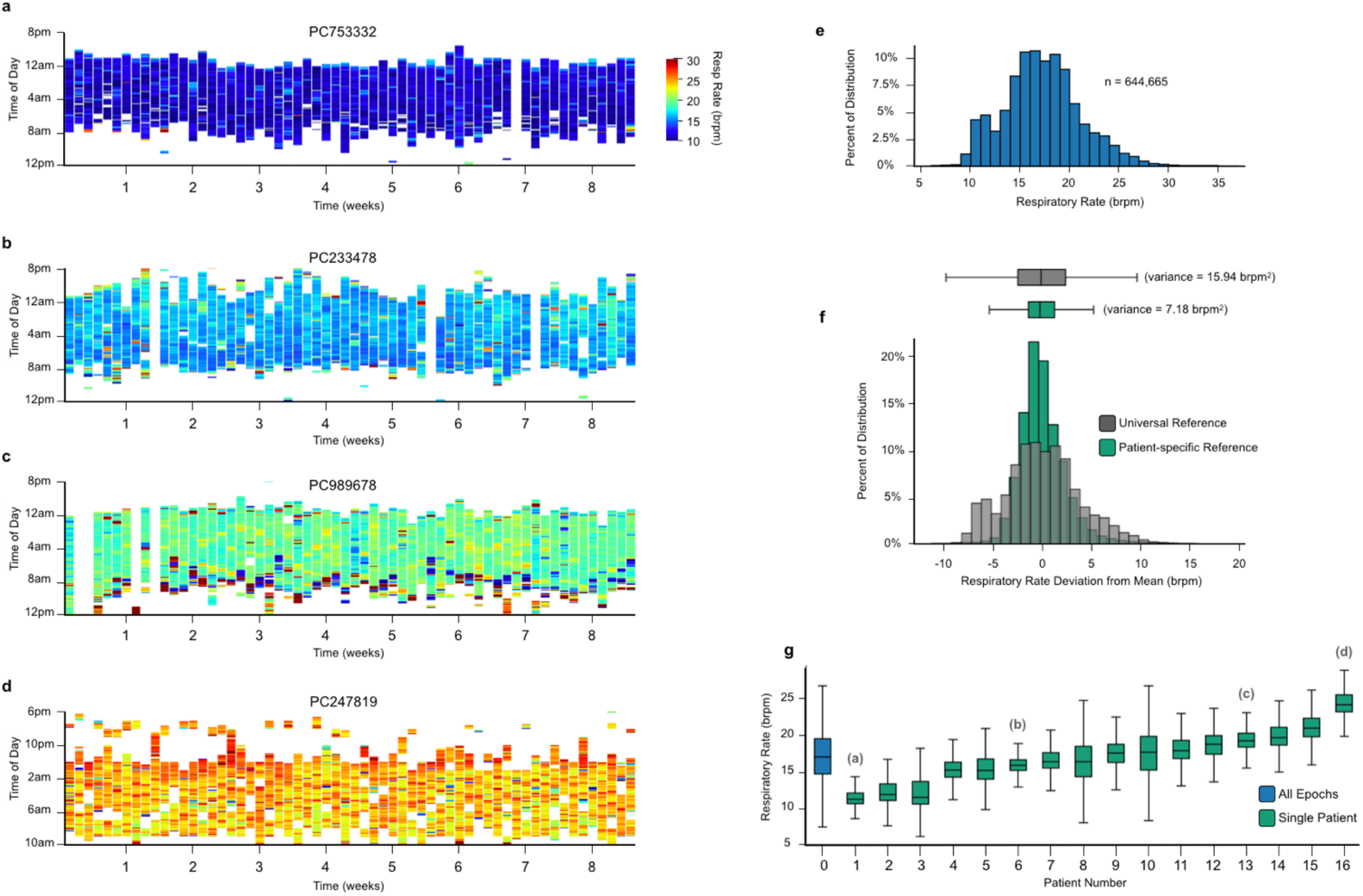
Nocturnal respiratory rate longitudinally in-home. **a-d**, 60-night heatmaps of 4 patients showing the intrapatient consistency and interpatient diversity of longitudinal respiratory rates in-home. Colorbar ranges from 10-30 brpm, values below 10 brpm not shown. **e**, Histogram of all epochs (n = 644,665) from the 16 stable patients with sufficient data (mean = 17.28 brpm, variance = 15.94 brpm^2^). **f**, Histograms of all epochs centered at the universal mean (gray) and each subject centered on their own overall mean (variance = 7.18 brpm^2^) (green). **g**, Box plot of the entire population as a single distribution (blue) and each individual patient (green). Labels in parenthesis highlight the patients shown in **a-d** with corresponding letter. For the box plots in **f** and **g**, the center line represents the median value, the boxes extend from the first to the third quartile, and the whiskers extend 1.5x the interquartile range before the first quartile and past the third quartile.

### Longitudinal NRR in patients with hospitalizations and clinical events

We next focused on patients who had clinical events during the monitoring period. During the 3400+ patient-nights of monitoring we observed 23 clinical events, defined as an overnight hospital stay, which is equivalent to a 30-day clinical event rate of ∼20%, consistent with the high-risk patient population that we enrolled. We observed hospitalizations for heart failure exacerbation (**Fig. 3a**), pneumonia responsive to antibiotics in a patient with heart failure on dialysis (**Fig. 3b**), failure to thrive in a diabetic man with chronic debilitating musculoskeletal pain (**Fig. 3c**), septic shock in a patient with long standing heart failure on dialysis (**Fig. 3d**), severe anemia from gastrointestinal bleeding in a patient with prior stroke (**Supplemental Fig. 6d**), as well as community-acquired COVID-19 infection in an immunosuppressed individual, hypoglycemia, hyperglycemia, localized cellulitis, and palpitations due to bigeminy. These clinical events occurred for diverse reasons, only some of which would be expected to have associated changes in respiratory rate.

**Figure 3.**
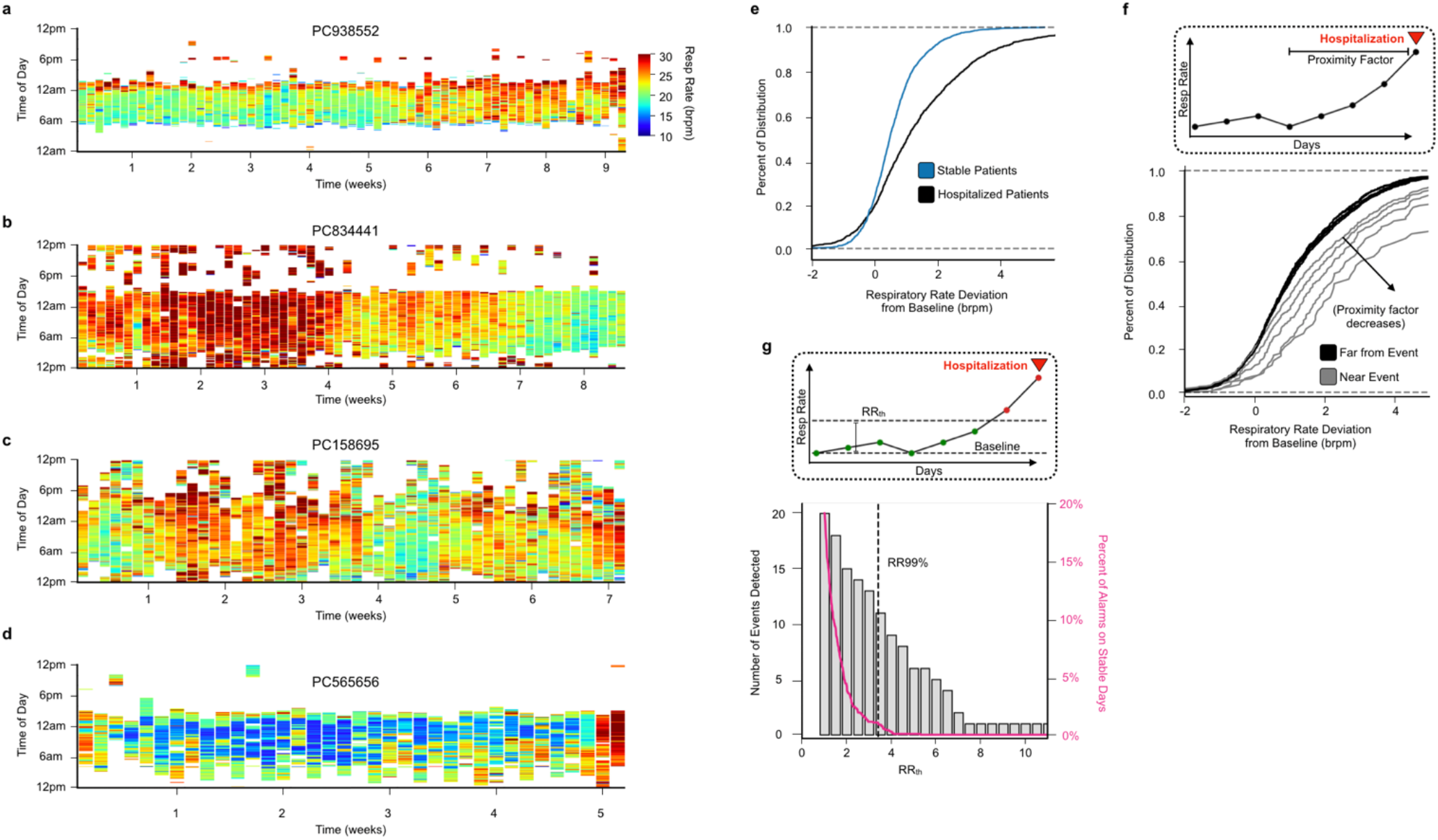
Comparison of stable and hospitalized patients. **a-d**, Heatmaps of hospitalized patients. Y-axis is time of day in hours. X-axis is time in weeks. Colorbar ranges from 10-30 brpm, values below 10 brpm not shown. **e**, CDF of the distribution of baseline deviations for stable patients (blue) and hospitalized patients (black). **f**, Inset shows that the Proximity Factor is a measure of how far a night is from a hospitalization event. Main plot shows a CDF of the distribution of baseline deviations for nights far from a hospitalization event (black), and for nights within 2, 5, 10, 20, and 30 nights from hospitalization event (gray). **g**, Inset shows that the RRth is a threshold with respect to baseline and that days below threshold are considered low risk (green) and days above are high risk (red). Main plot shows the number of hospitalization events detected (gray bars, left y-axis) and the number of days among stable patients that were labeled “high risk” (pink line, right y-axis) for varying values of RRth. RR99% (black dashed line) is a value of RRth where 99% of stable days are considered low risk.

To analyze the quantitative differences between hospitalized and non-hospitalized patients, we first defined a causal patient-specific baseline as the lowest sustained NRR (see methods). We then plotted the deviation of NRR compared to baseline for each patient type. The hospitalized group had a 4-fold greater variance compared to the stable group (3.78 brpm^2^ and 0.84 brpm^2^ respectively), the KS statistic for the two groups had a p value <0.0001 indicating that the two distributions were significantly different, and by inspection of the CDF the distinction between the two groups is due to the hospitalized group having a greater proportion of elevations from baseline (**Fig. 3e**). We next asked whether the differences were concentrated and localized to days immediately preceding the hospitalization event. We defined a proximity factor to specify windows in advance of hospitalization for analysis. As the proximity factor decreased, the deviation from baseline increased suggesting that it may be possible to detect impending hospitalization by longitudinally monitoring deviations of NRR from baseline (**Fig. 3f**). We set a threshold NRR at a defined magnitude (NRRth) above the patient-specific baseline. When patients exceeded the threshold, they were classified as “in-alarm” (red) while those who remained below threshold were classified as “not in-alarm” (green) (**Fig. 3g**). The optimal value of NRRth can then be selected to balance detection of hospitalizations (true positives) while minimizing alarms for patients who did not ultimately require hospitalization (**Fig. 3g**). We did not classify the latter as “false alarms” since in some cases, these appeared to be subclinical events with a gradual rise-and-fall pattern that did not require hospitalization but nevertheless represented physiologic deviations that would likely benefit from outreach (**Supplemental Fig. 5**).

### Development of an impending hospitalization

To evaluate performance of this approach, we selected a NRRth threshold value to limit above-threshold days to <1% in non-hospitalized patients (NRR99%) and applied it to hospitalized and non-hospitalized patients **(Fig. 4)**. This resulted in 2 alarm regions in 2 non-hospitalized patients over 1707 days of monitoring, one of which had a characteristic rise-and-fall pattern suggestive of a subclinical event **(Supplemental Fig. 5)**. By comparison, we alarmed on 11 of 23 hospitalizations, which occurred for reasons including heart failure with volume overload, anemia requiring transfusion, pneumonia, acute COVID-19 infection, and septic shock. To evaluate the window of early detection, we aligned the hospitalizations to compare the duration of each prodrome. The longest prodrome (∼3 weeks) was associated with a volume overload event from heart failure exacerbation and 2 of the shortest prodromes (∼1 day) were associated with infections. Such warning periods would provide ample time for patient calls, home visits, urgent clinic visits, and escalation of diuretics. Meanwhile, although infections may be associated with shorter prodromes, the ability to detect deterioration at home before patients self-reported symptoms and elected to self-present for clinical care could be very useful, since, for example, every hour of delay in initiating antibiotics is associated with increased mortality in sepsis.

**Figure 4.**
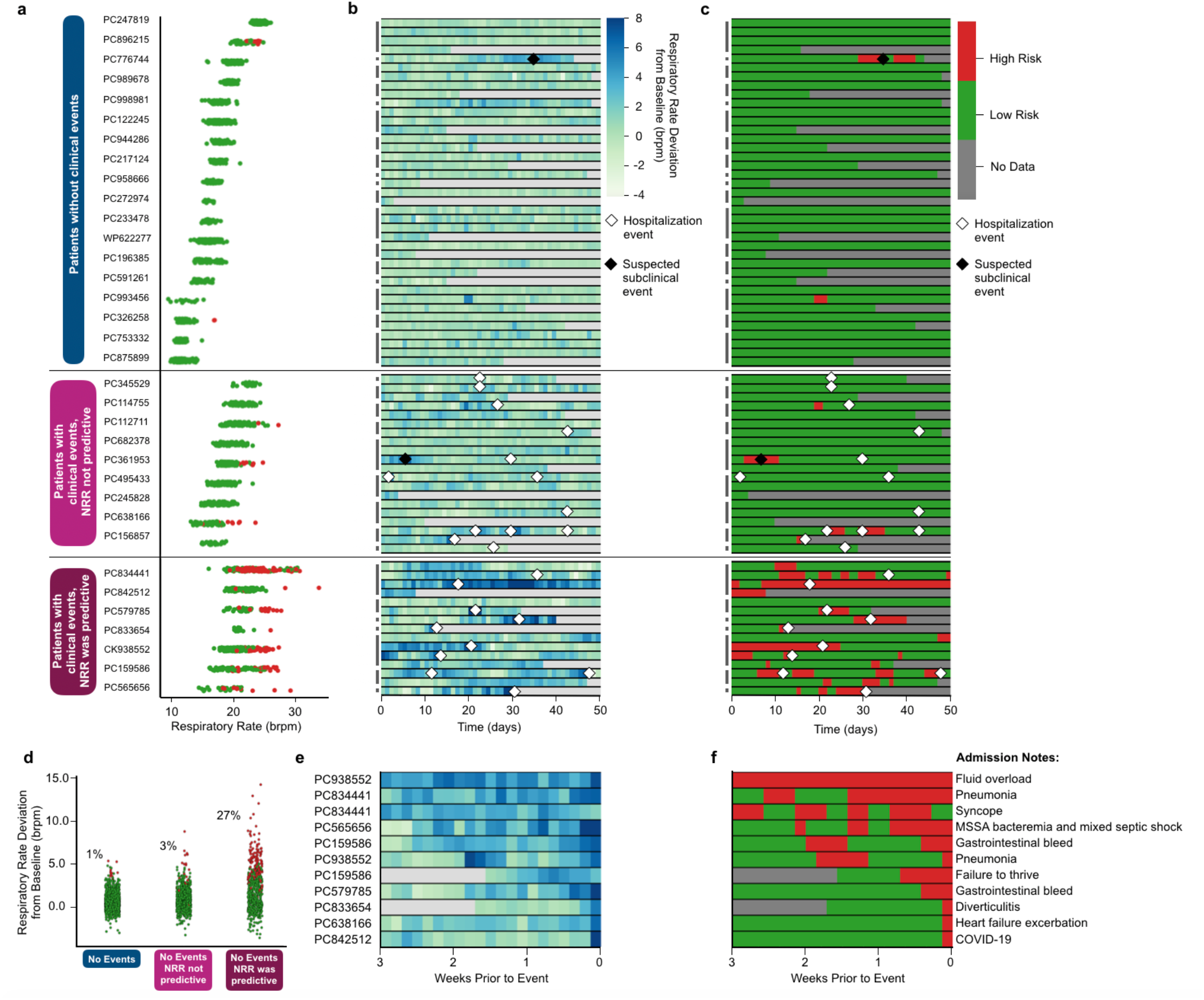
All patient-nights with risk assessment. **a**, Dot plot of nocturnal respiratory rate for all nights of patient data, colored by low risk (green) and high risk (red). Patients are grouped into 3 categories, patients who had no clinical events (blue), patients who had clinical events but for whom NRR was not predictive (light magenta), and patients who had clinical events and for whom NRR was predictive (dark magenta). **b-c**, Heatmaps of respiratory rate deviation from baseline **(b)** and risk assessment **(c)** for all nights of patient data. Patients may span multiple rows. Gray used for padding. Hospitalization events annotated with white diamond. Suspected sub-clinical events annotated with gray diamond. Days with less than 3 hours of epochs not shown. **d**, Dot plot of NRR deviation from baseline for all patient-nights. Colored by low risk (green) and high risk (red) and grouped into the same 3 groups described above. **e-f**, Heatmaps of NRR deviation from baseline **(e)** and risk assessment **(f)** for 3 weeks prior to hospitalization for events with respiratory rate prodromes. Days with less than 3 hours of epochs not shown. The same colorbars from **b-c** are used in **e-f**.

## DISCUSSION

Respiratory rate is a fundamental vital sign that is often clinically neglected, except at its extremes. Here, we show that the nocturnal respiratory rate (NRR) is a powerful biomarker that can be measured longitudinally and in an adherence-independent manner without requiring any patient engagement. Across 2000 patients from the Sleep Heart Health Study and 3400 patient-nights of longitudinal home bed monitoring in high-risk patients, we discovered patterns of clinical stability and prodromes of impending hospitalizations for diverse causes that are amenable to early automated detection. During periods of clinical stability, each patient exhibited low night-to-night variability compared to higher interpatient differences. This argues in favor of using patient-specific referenced analysis rather than traditional cross-sectional analysis. Doing so enables early detection of respiratory changes days and weeks prior to hospitalization for conditions such as heart failure, diverticulitis, severe anemia due to gastrointestinal bleeding, pneumonia, acute COVID-19 infection, and septic shock. Although the number of events was small, we believe that in general, longer prodromes will be associated with fluid overload while shorter prodromes will be associated with infections.

As expected, not all causes of hospitalizations were preceded by respiratory prodromes. For example, mechanical falls, localized skin and soft tissue infections, some hypo/hyperglycemic episodes and some arrhythmias did not alter NRR. As such, we do not recommend interpreting performance as a diagnostic test for all-cause hospitalizations using a receiver operator curve quality metric. This is in part because ground truth is not well defined, as false positives may represent subclinical events that would benefit from outreach, and because false negatives may simply represent hospitalizations for which there is no expected respiratory prodrome. Instead, our goal is to maximize early identification of clinical events that outperforms patient-reported symptoms while minimizing alarms that, in retrospect, are felt to have no clinical correlate.

Because the monitoring is automated and because patient heatmaps place each day’s data in the context of all prior days, one can review large cohorts via a web-based dashboard daily and confirm NRR stability in seconds for each patient, leaving ample time to more closely analyze and follow-up with patients who depart from their baseline. For example, consider a care coordinator (CC) who is responsible for 200 high-risk patients with an average hospitalization rate of 1.5 per year. The CC would expect an average of ∼1 hospitalization per business day. Since it is unknown which patients are clinically deteriorating on any given day, a common strategy is to schedule and perform routine structured phone calls (e.g., 1 per month, which for 200 patients, totals ∼10 calls per business day with an interval of ∼30 days between calls). Contrast that with a workflow based on automated remote monitoring and NRR-prompted calls. A CC can review all 200 patients every day in less than 10 minutes leaving the remainder to focus on phone-calls, in-home visits, and unscheduled clinic visits, for the ∼5-10 patients per week who are becoming unstable. This provides 5-10x more time for extensive discussions with the patients and family, home visits, targeted examination and diagnostic testing, coordination of care, and if necessary, interventions. Despite a 20% 30-day event rate, the number of patients with false alarms was low, and among those, a few were likely subclinical events for which outreach would have been warranted (2 of which are shown in Supplemental Fig. 5). Therefore, we have not found unwarranted alarms to be frequent or burdensome. Use of automated NRR-based early warning has potential to enable efficient allocation of care coordinator resources.

NRR is also an attractive biomarker because it can be reliably monitored without biasing towards technophiles or highly engaged patients. The patients in our study were primarily elderly; many lived alone; most had multiple chronic diseases, several were blind, frail, or bed-bound; yet we were able to record NRR longitudinally for more than 200 nights (the fully duration of our study). Beyond the current analysis, we have continued monitoring individuals from this cohort for over 1.5 years. Most of the patients in our cohort were part of a university population health program that employs care coordinators and a home health team to monitor and care for clinical deteriorations in the outpatient setting. Additionally, NRR is an attractive biomarker because it can be measured by many different sensors. Adherence-independent strategies can be based on strain gauges placed under the bed *(20)*, mattress sensors *(21-24)*, or ultrawideband radar systems *(25, 26)*. For highly engaged patients, there are many wearables and smartphone apps capable of measuring respiratory rates *(27-29)*. NRR can also be measured via CPAP ventilatory support *(30)* and implanted intracardiac devices *(31-34)*.

What should be done in response to early recognition? Management of clinically dynamic patients in the outpatient setting is a largely unstructured domain of outpatient medicine that is likely to mature as early detection technologies continue to be developed. Currently, we use early recognition of impending hospitalizations to prompt patient phone calls and, if necessary, intervention or escalation of care. Future work will test whether communicating NRR to care coordinators accompanied by structured workflows can be prospectively tested to determine whether early detection and intervention improves outcomes. This is particularly promising because many causes of increased NRR have established interventions. For example, diuretics can be uptitrated for fluid overload in heart failure or chronic kidney disease while antibiotics can be started for localized infections or combined with steroids for COPD exacerbations. For more severe deteriorations, early evaluation in the ER and consideration of hospitalizations may reduce overall length of stays, need for ICU, or improve clinical outcomes after discharge. These questions will need to be tested prospectively to determine if NRR-guided care can improve the quality and cost of care as well as the quality of life for patients and their families.

## CONCLUSION

In summary, we show that NRR, when monitored longitudinally using non-contact adherence-independent bed sensors, is a promising biomarker of impending hospitalizations. The potential for early recognition of acute illnesses makes outpatient chronic disease management a data-driven science and in doing so, achieves the triple aim of improving patient satisfaction, improving quality of care and access for populations all while reducing health care costs *(35)*.

## Data Availability

All epoch data produced in the present study are available upon reasonable request tot he authors.

## Acknowledgements

We thank Mr. Mike McCauley for the BCM 2835 library used on the respiratory rate sensor. The work was funded by UCOP TRDRP Award, and NIH-NINR R21NR018558 (K.R.K.). The Sleep Heart Health Study (SHHS) was supported by National Heart, Lung, and Blood Institute cooperative agreements U01HL53916 (University of California, Davis), U01HL53931 (New York University), U01HL53934 (University of Minnesota), U01HL53937 and U01HL64360 (Johns Hopkins University), U01HL53938 (University of Arizona), U01HL53940 (University of Washington), U01HL53941 (Boston University), and U01HL63463 (Case Western Reserve University). The National Sleep Research Resource was supported by the National Heart, Lung, and Blood Institute (R24 HL114473, 75N92019R002).

## Author Contributions

N.H. and K.R.K. designed and conducted all aspects of the project and wrote the manuscript. A.W. consented patients, installed sensors, and performed data transfer troubleshooting. D.T.B. and Q.M.B. enrolled patients and analyzed clinical histories. S.K., R.B.R., and K.S. analyzed clinical histories. R.L.O. oversaw sensor validation studies. R.L.O., P.A., and K.R.K. designed and interpreted the clinical study. All authors analyzed data and edited the manuscript.

## METHODS

### Sleep study analysis

The subjects for our sleep study analysis were taken from a 2,400-subject subset of the Sleep Heart Health Study conducted by the National Heart Lung & Blood Institute **(Fig. 1)**. Subjects wore a chest belt during the night sampled at 10 Hz with a 0.05 Hz hardware high-pass filter. To calculate the NRR, we first took the signal from the chest belt of each subject and smoothed it with a moving mean filter (window size of 3 samples). Because the chest belt signal saturated, to isolate regions of probable movement, we then multiplied regions near the saturation point by a factor of 10 and applied a moving variance filter (window size of 5 seconds) to differentiate regions of low amplitude steady signal from high amplitude signal. We required regions of steady physiology to be at least 10 seconds in duration. Peak-finding was then performed on the resultant regions of steady physiology. The respiratory rate was estimated based on the median inter-breath interval within a moving window (shift of 30 seconds, window size of 5 minutes). To reject regions that were noise rather than physiology, we performed autocorrelation analysis as an alternative method of calculating the respiratory rate and required the result to be within a physiologically reasonable range (<40 bpm). NRR epochs were rejected if they were not within a physiologically reasonable range (6 brpm<x<40 brpm). Subjects were required to have at least 3 hours of quality epochs during the night to be included. The 2000 subject heatmap was sorted by mean respiratory rate. The x-axis in Figure 1b-c is cropped such that all bins with >0.01% of data are shown. Variance comparison analysis was calculated using a Levene test implemented by scipy.stats.levene. Calculation of skew used the Fisher-Pearson coefficient of skewness and was implemented by scipy.stats.skew.

### In-home study

We started with a cohort of 41 in-home patients. We excluded patients who had less than 5 nights of data (impacting 6 patients) and we excluded an additional patient for technical reasons (see discussion of two-person issue below). Two patients (PC196385 and PC842512) had gaps in their records longer than 2 months and regions these were not included in our accounting of nights monitored or nights missed. Two patients (PC938552 and PC156895) had 5 nights with timing mismatch at the beginning of their records and these nights were not included. Two nights from PC345529 and PC989678 and 4 nights from PC217124 were excluded due to artifacts. Eight nights from PC245828 were excluded, 3 for artifacts, and 5 because, based on weight analysis, the data on those nights was not from our patient. Two patients (PC245828 and PC834441) had one set of sensors under their bed and another set of sensors under their recliner and these 2 data streams were combined to create a composite record for each of them. The first 16 nights of PC233478 had a large proportion of artifacts and required a reinstall so we started their record after the reinstall. From the beginning to end of each patient’s record there were 3403 nights of data total and 3056 nights with a sufficient number of epochs. On average, 91% of each patient’s record contained nights with a sufficient number of epochs (**Supplemental Fig. 2**).

Nine of our patients share a bed with a partner and so for these datasets there is uncertainty as to which individual is contributing to the NRR measurement from our sensor (one patient was excluded from our study due to this issue). Of the 9 that were included, 1 patient (PC196385) was able to be partially isolated based upon weight and timing. Additionally, this patient had 2 hospitalization events that were successfully detected, indicating that this data set is matched to the correct person. For the remaining 8 patients we assumed that we are monitoring the correct individual and future work will employ two sensors to disambiguate individuals who share a bed. Six of these 8 patients (PC217124, PC326258, PC753332, PC938552, PC944286, and PC989678) were not hospitalized which matches our NRR assessment as these individuals had almost entirely low risk nights. We acknowledge the possibility for false negatives for these patients if their data was reflective of the patient’s partner and the partner was hospitalized for a disease exacerbation during our monitoring period. Two patients (PC361953 and PC682378) had events that went undetected by NRR and possibly these false positives were caused by this two-person issue.

To monitor the NRR, we used BedScales, a non-contact adherence independent sensor that is placed under the legs of the bed and transmits data every hour to Amazon Web Services cloud storage where it is processed in real-time through a pipeline of analytics that extract the respiratory rate *(20)*. The processing pipeline receives a signal from the sensors under each bed leg, performs bandpass frequency dependent filtering and combines all individual sensor signals into a composite signal using Principle Component Analysis. A moving variance filter (window size of 5 seconds) was used to isolate regions of steady physiology (these regions were required to be at least 10 seconds in length) and a weight-based method was primarily used to determine times when the patient is in bed. Peak-finding is then performed on the resultant regions of steady physiology. The respiratory rate is estimated based on the median inter-breath interval within a moving window (shift of 30 seconds, window size dilates to capture at least 5 minutes of steady breathing). To reduce artifacts, some patient-nights were processed through a slightly different pipeline that detected if the patient was in bed using a different weight-based method, that found steady regions on a signal filtered using a 2^nd^ order bandpass with a slightly lower upper cutoff (1 Hz instead of 1.5 Hz), that removed inter-breath intervals less than 1.5s or intervals that spanned a movement from consideration, and that used a fixed window size of 5 minutes rather than a dilating window. The resulting respiratory rate epochs from the pipelines were required to be within a physiologically reasonable range (6<x<40) and nights were required to contain at least 3 hours of epochs to be included. By convention, our “patient-nights” are defined as running from 12pm-12pm (changes due to daylight savings are not considered). Using the EMR, we recorded the significant interactions that our in-home patients had with clinicians. We separated the patients into a stable cohort (18 patients) and a hospitalized cohort (16 patients). We included in the hospitalized cohort all patients who were admitted overnight except for two patients (PC993456 and WP62227) because they were hospitalized for falls rather than a disease exacerbation. We included as an admission one patient (PC158695) who was admitted to a Skilled Nursing Facility. For correlating our in-home data with hospitalization events, standard calendar days were used and so events were recorded if they occurred up to midnight during the final night of monitoring for each individual.

### Longitudinal in-home analysis

To assess the longitudinal stability of nocturnal respiratory rate for in-home patients (**Fig. 2**), we chose the patients who were not admitted to the hospital for a disease exacerbation and who had at least 30 nights of data, resulting in 480 nights total from 16 patients. The epochs for assessing stability were taken from a window of 30 nights that began at night 1 for these patients. For patients who had large gaps (>14 nights) of missing data during this period, the 30-night window started after the gap period (this impacted patients PC196385 and PC217124). The x-axis in Figure 2b-c is cropped such that all bins with >0.01% of data are shown. Variance comparison analysis was calculated using Levene test implemented by scipy.stats.levene.

### Baseline calculation and self-referencing

To derive a baseline nocturnal respiratory rate for each patient, we found the causal moving average of each night with a window size of 5 nights and then took the rolling minimum of this value as it evolved across time. This ensured that each baseline value was the result of multiple nights of monitoring and that if our sensors were installed to a patient who was actively returning to their normal range, the derived baseline value would gradually decrease as the individual’s respiratory rate returned to normal (**Supplemental Fig. 3**). Since there is no causal moving average defined for the first x-1 nights where x is the window size, when the baseline was subtracted from the nightly NRR estimate, all of the nights prior to a genuine baseline measure were set to a null value and not considered in subsequent analysis. A maximum of 1 night of missing data was allowed within the window of nights that was used in setting the moving average.

### Clinically stable versus hospitalized patients

To see if there is any difference in nocturnal respiratory rate between clinically stable and hospitalized patients (**Fig. 3e**), we compared the distributions of baseline deviations across all nights for these two populations. To compare distributions comprised of nights near and far from hospitalization events (**Fig. 3f**), we defined a Proximity Factor that defined how many nights prior to the event were placed into the “near” distribution. To take into consideration that after the hospitalization event, the patient may be fully recovered, or may still be sick and recovering, we set a buffer value of 7 nights after the event and those nights were not placed into either the “near” or “far” distributions. To compare the CDFs of the distributions in Figure 3e-f, we used a Kolmogorov-Smirnov test for goodness of fit, implemented by scipy.stats.kstest.

For our risk assessment strategy, we set a threshold (RRth) that is a specified amount above baseline and in general, all nights above the threshold were considered high risk nights. To reduce noise from artifacts or from sporadic high-risk alerts, we incorporated 3 other considerations (**Supplemental Fig. 4**): (i) A night must either be the second consecutive above RRth to be considered high risk or (ii) the night must be above a higher threshold, RRth+ to be considered high risk. (iii) Nights that dropped below RRth could still be considered high risk if that night, and the other consecutive preceding high-risk nights (if any) had a mean that was still above the threshold. Considerations (i) and (ii) prevent sporadic or errant nights from being labeled high risk but allow for detection of acute conditions that suddenly raise the NRR (**Supplemental Fig. 7c**). Based on our dataset, we set RRth+ to be 1.75 brpm above RRth. Consideration (iii) is a causal way to prevent high risk regions from being fractured by nights that dip slightly below the threshold.

To count the number of events that were preceded by a rise in NRR (**Fig. 3g**), we set a margin value of 3, and any event that occurred within 3 nights of a high-risk night was considered to have been detected by NRR analysis. We set RRth based upon the value that would yield 1% or fewer high-risk nights in patients that were clinically stable (RRth99%). For our dataset, this value was 3.41 brpm.

### Risk assessment

We applied our risk assessment strategy to the entire dataset (**Fig. 4**). We grouped patients into three categories (i) patients who had no hospitalization events (ii) patients who were hospitalized but for whom NRR was not predictive and (iii) patient who were hospitalized and for whom NRR was predictive. To create the heatmaps in Figures 4b-c we arranged all patient-nights in columns of 50 nights. Some patients spanned multiple rows and gray was used for padding to keep the columns even. Nights without data are not shown. We aligned all hospitalization events together and displayed 21 nights prior to each event (**Fig. 4f**). One patient in this figure, (PC842512) had their high-risk night on the same day as his admission so this was adjusted for display purposes.

## SUPPLEMENTAL FIGURES

**Supplemental Figure 1.**
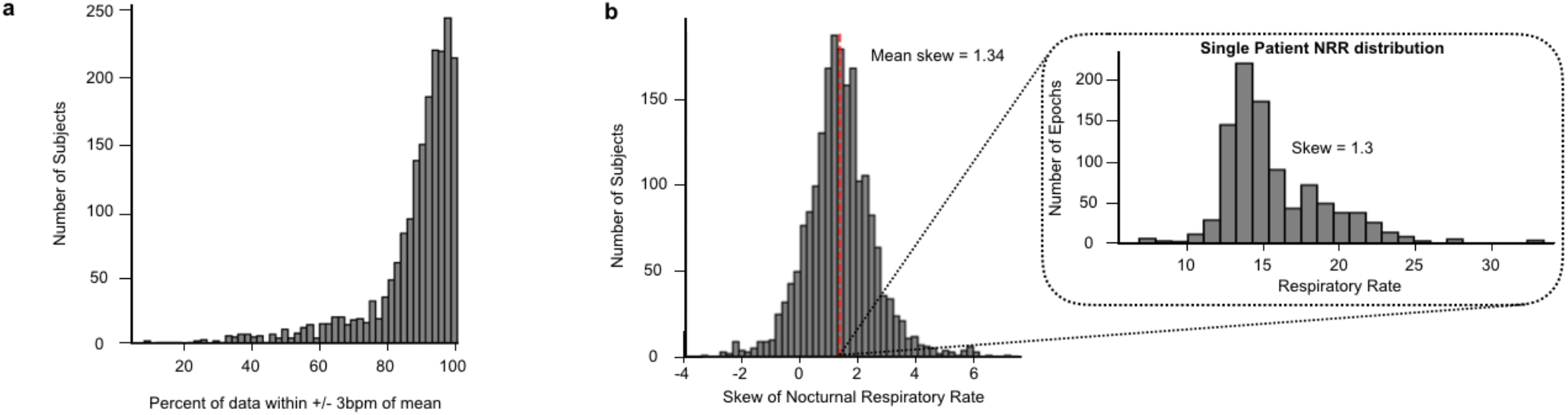
Nocturnal respiratory rate sleep study intra-night statistics. **a**, Histogram of percent of respiratory rates within 3 brpm of each sleep study subject’s mean. **b**, Histogram of skew per sleep study subject. Red dashed line indicates mean of skew (1.33). Inset show nocturnal respiratory rate distribution for a patient with a typical skew value (1.3).

**Supplemental Figure 2.**
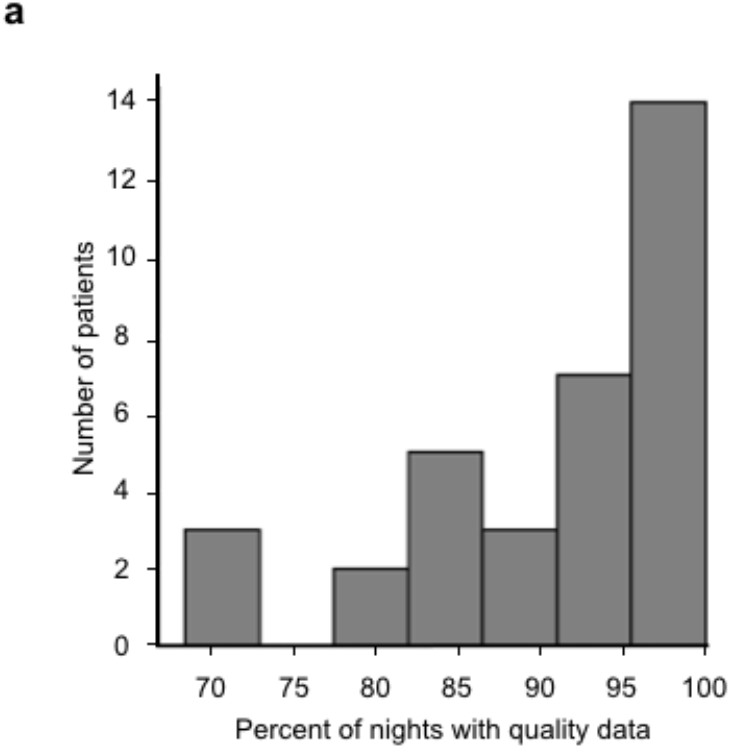
Percent of nights with sufficient data. **a**, Histogram of percent of nights within monitoring period that contained at least 3 hours of quality epochs.

**Supplemental Figure 3.**
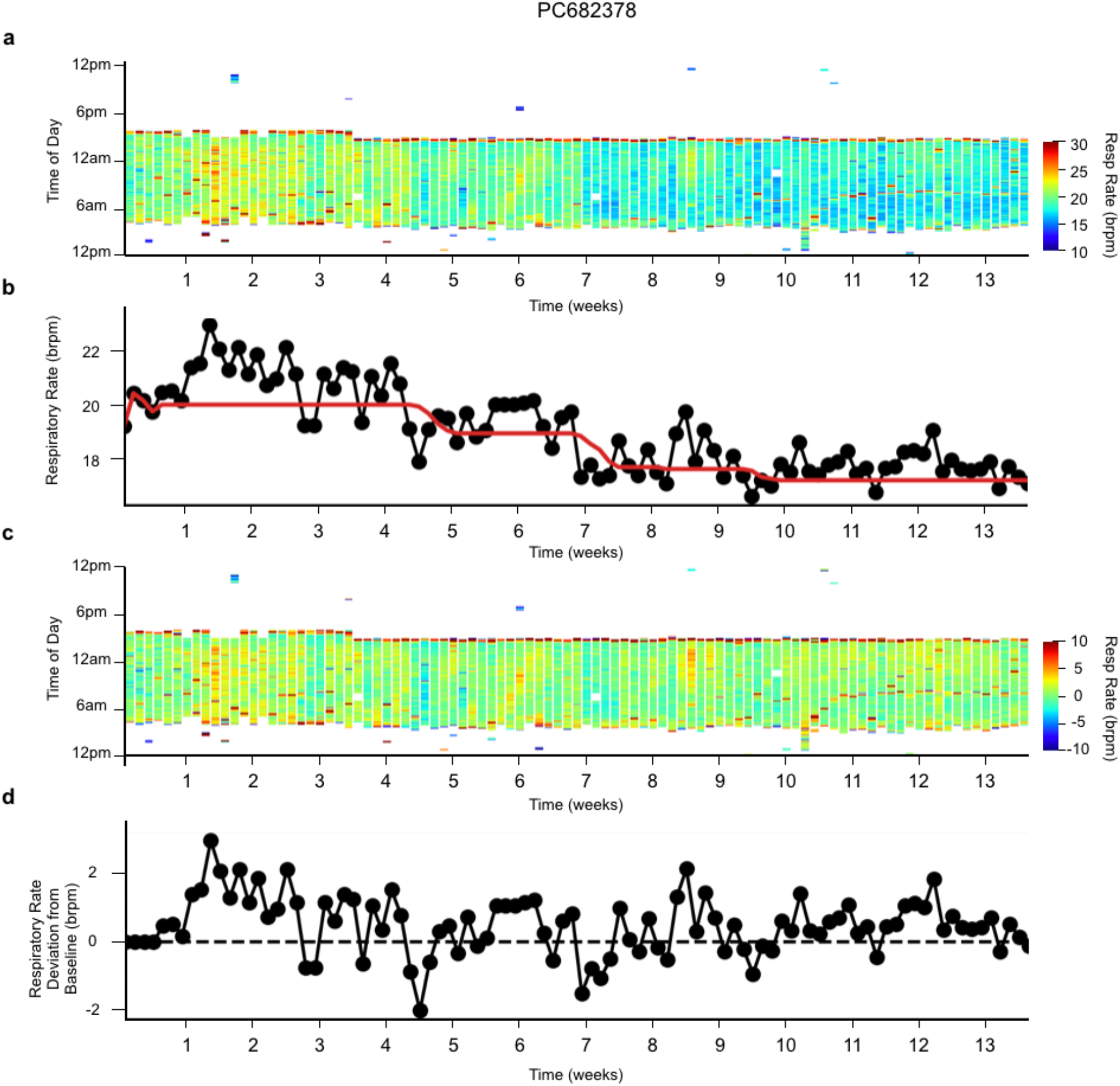
Causal baseline. **a**, Heatmap of NRR for a patient with a monotonically decreasing brpm. Colorbar ranges from 10-30 brpm, values below 10 brpm not shown. **b**, Nightly median respiratory rate (black) with causal baseline annotated (red). **c**, Heatmap of NRR with baseline subtracted. Colorbar ranges from -10 to 10 brpm, values below -10 not shown. **d**, Nightly median respiratory rate (black) with baseline subtracted.

**Supplemental Figure 4.**
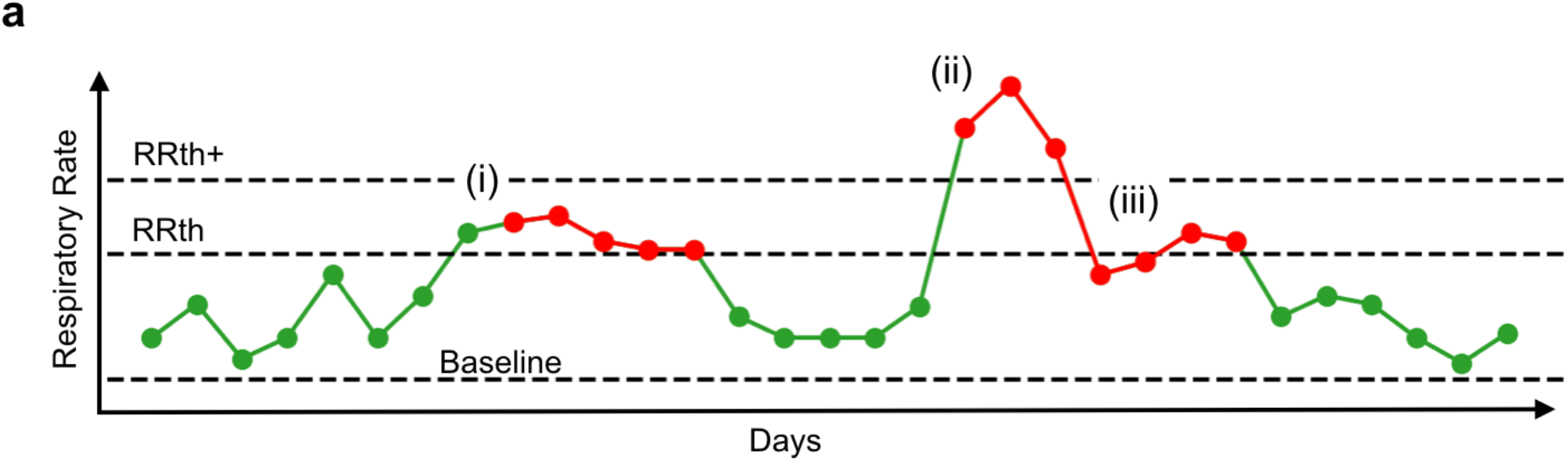
Risk assessment strategy. **a**, Illustration of respiratory rates across time colored by low risk (green) and high risk (red). Horizontal dashed lines indicate the baseline NRR, the NRR threshold (RRth) and the additional NRR threshold (NRR+). In general, points below the first threshold (RRth) are low risk and points above are high risk. Roman numerals highlight regions of special consideration (see Methods). **(i)** Demonstrates that nights above the first threshold are only considered high risk if they are the second consecutive point above the threshold. **(ii)** Demonstrates that if a night is above the second threshold (NRR+) it is always considered high risk. **(iii)** Demonstrates that nights below the first threshold are still considered high risk if the days preceding them have an overall mean that is above the first threshold.

**Supplemental Figure 5.**
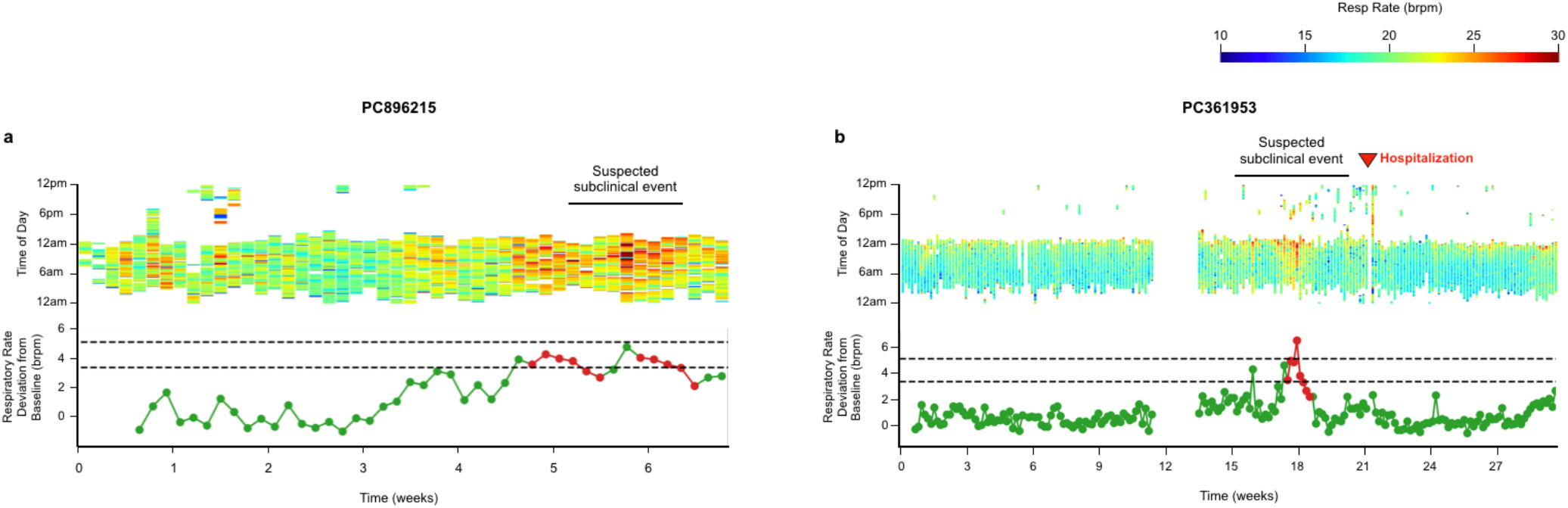
Suspected subclinical events. Heatmap and corresponding NRR deviation from baseline for two patients who both exhibited a steady rise and fall of brpm indicative of a disease exacerbation. Heatmap colorbar ranges from 10-30 brpm, values below 10 brpm not shown. NRR excursions colored by low risk (green) and high risk (red). Black dashed lines show RRth (bottom) and RRth+ (top). PC896215 (a) was never seen in clinic during our monitoring period and PC361953 (b) was seen in clinic for orthostatic hypotension 3 weeks after the suspected subclinical event was resolved.

**Supplemental Figure 6.**
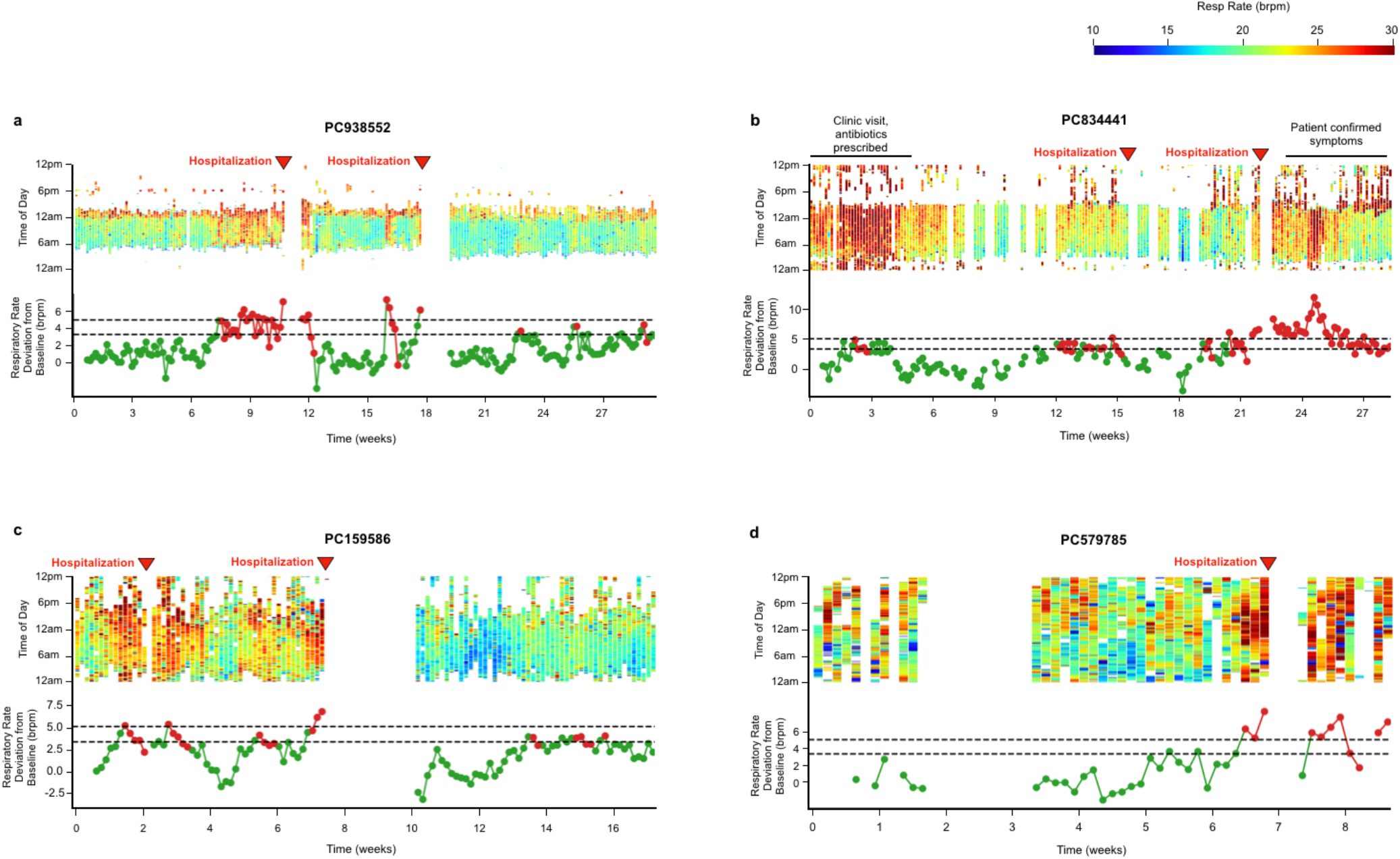
Heatmaps for patients with clinical events, NRR predictive. Heatmap and corresponding NRR deviation from baseline for 4 patients whose events were preceded by a rise in NRR. Heatmap colorbar ranges from 10-30 brpm, values below 10 brpm not shown. NRR excursions colored by low risk (green) and high risk (red). Black dashed lines show RRth (bottom) and RRth+ (top). **a**, Patient had two hospitalization events, the first for fluid overload (+/- pneumonia), the second for pneumonia and mild fluid overload. **b**, Patient had two hospitalization events, the first for syncope, the second for pneumonia and mild fluid overload. **c**, Patient had two clinical events, the first was a hospitalization for a gastrointestinal bleed, the second was a referral to a skilled nursing facility for failure to thrive (inability to complete ADLS and for knee pain). **d**, Patient was hospitalized due to concern for gastrointestinal bleed.

**Supplemental Figure 7.**
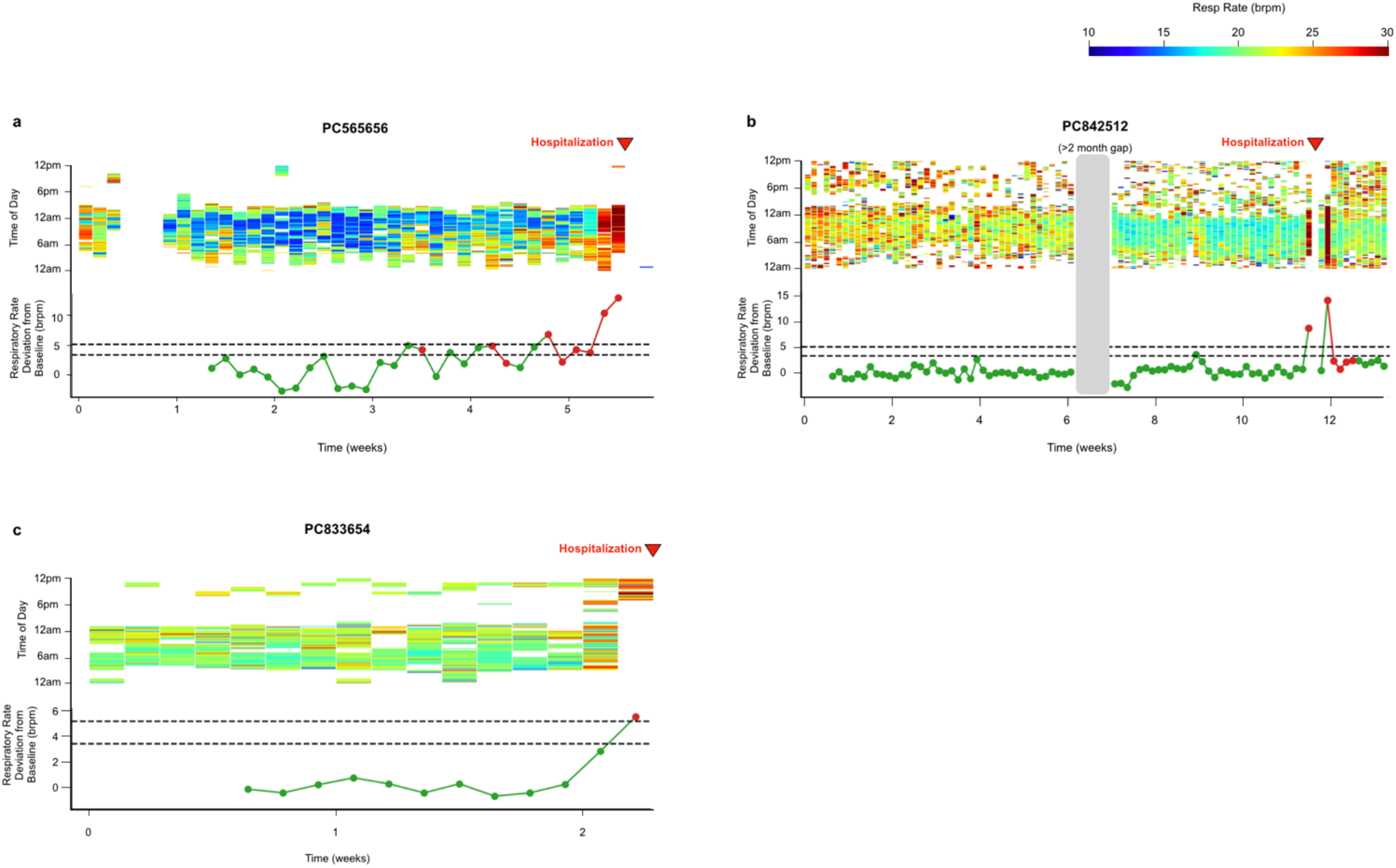
Heatmaps for patients with hospitalization events, NRR predictive. Heatmap and corresponding NRR deviation from baseline for 3 patients whose events were preceded by a rise in NRR. Heatmap colorbar ranges from 10-30 brpm, values below 10 brpm not shown. NRR excursions colored by low risk (green) and high risk (red). Black dashed lines show RRth (bottom) and RRth+ (top). **a**, Patient was referred to emergency department for shortness of breath and difficulty walking. Was admitted for MSSA bacteremia and mixed septic shock. **b**, Patient was hospitalized for fever and chills. Was found to have COVID-19. **c**, Patient was seen in emergency department for altered mental status and malaise. Was hospitalized for diverticulitis.

**Supplemental Figure 8.**
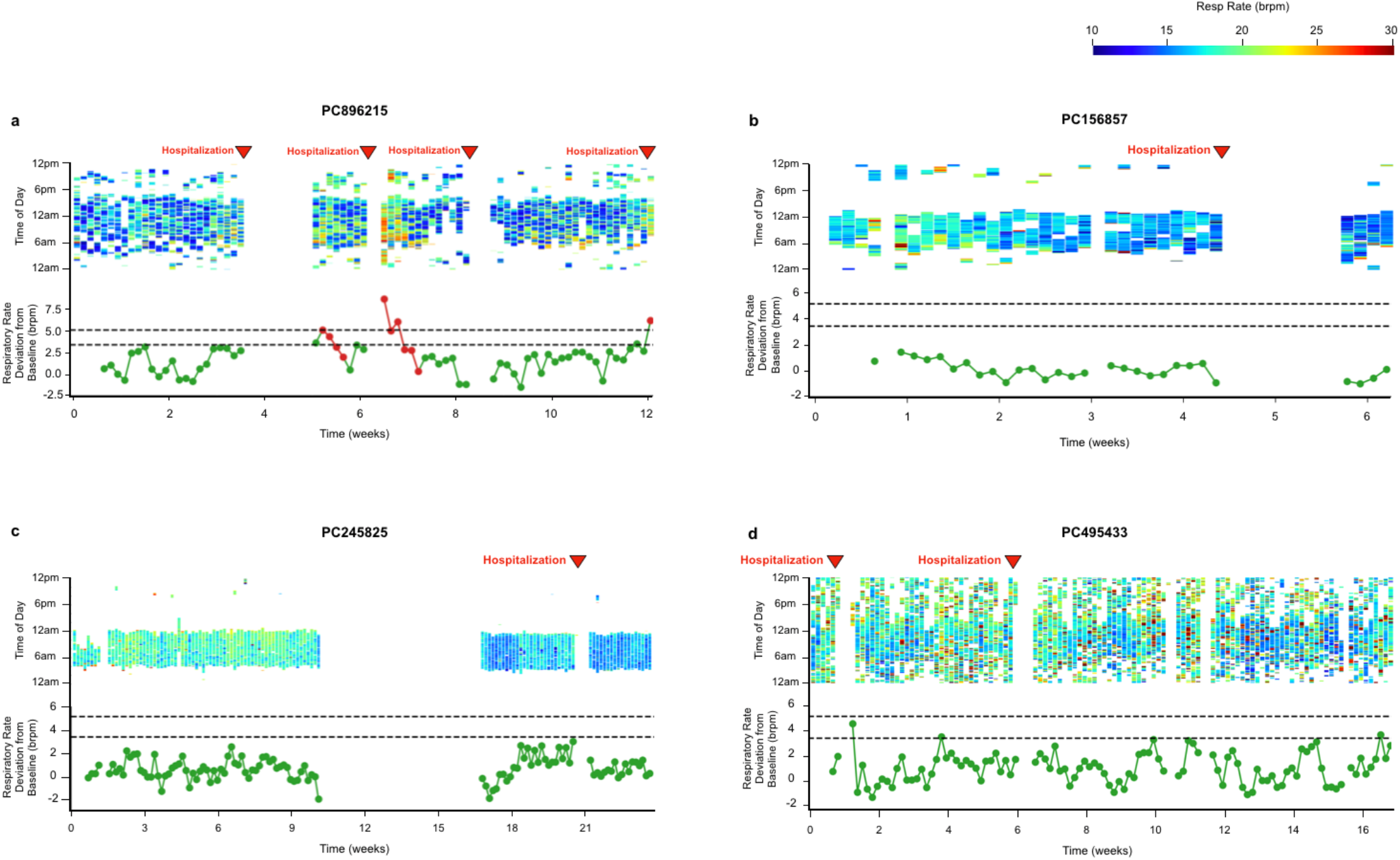
Heatmaps for patients with hospitalization events, NRR not predictive. Heatmap and corresponding NRR deviation from baseline for 4 patients whose hospitalization events were not preceded by a rise in NRR. Heatmap colorbar ranges from 10-30 brpm, values below 10 brpm not shown. NRR excursions colored by low risk (green) and high risk (red). Black dashed lines show RRth (bottom) and RRth+ (top). **a**, Patient had 4 hospitalization events, the first for volume overload, the second for symptomatic bigeminy and ectopy, the third for cellulitis, and the fourth for heart failure exacerbation (this event was associated with a rise in NRR). **b**, Patient was referred for cellulitis symptoms, was found to have heart failure. **c**, Patient was hospitalized for congestive heart failure exacerbation. **d**, Patient had two hospitalization events, the first for over-diuresis, the second for hyperglycemia and hypovolemia.

**Supplemental Figure 9.**
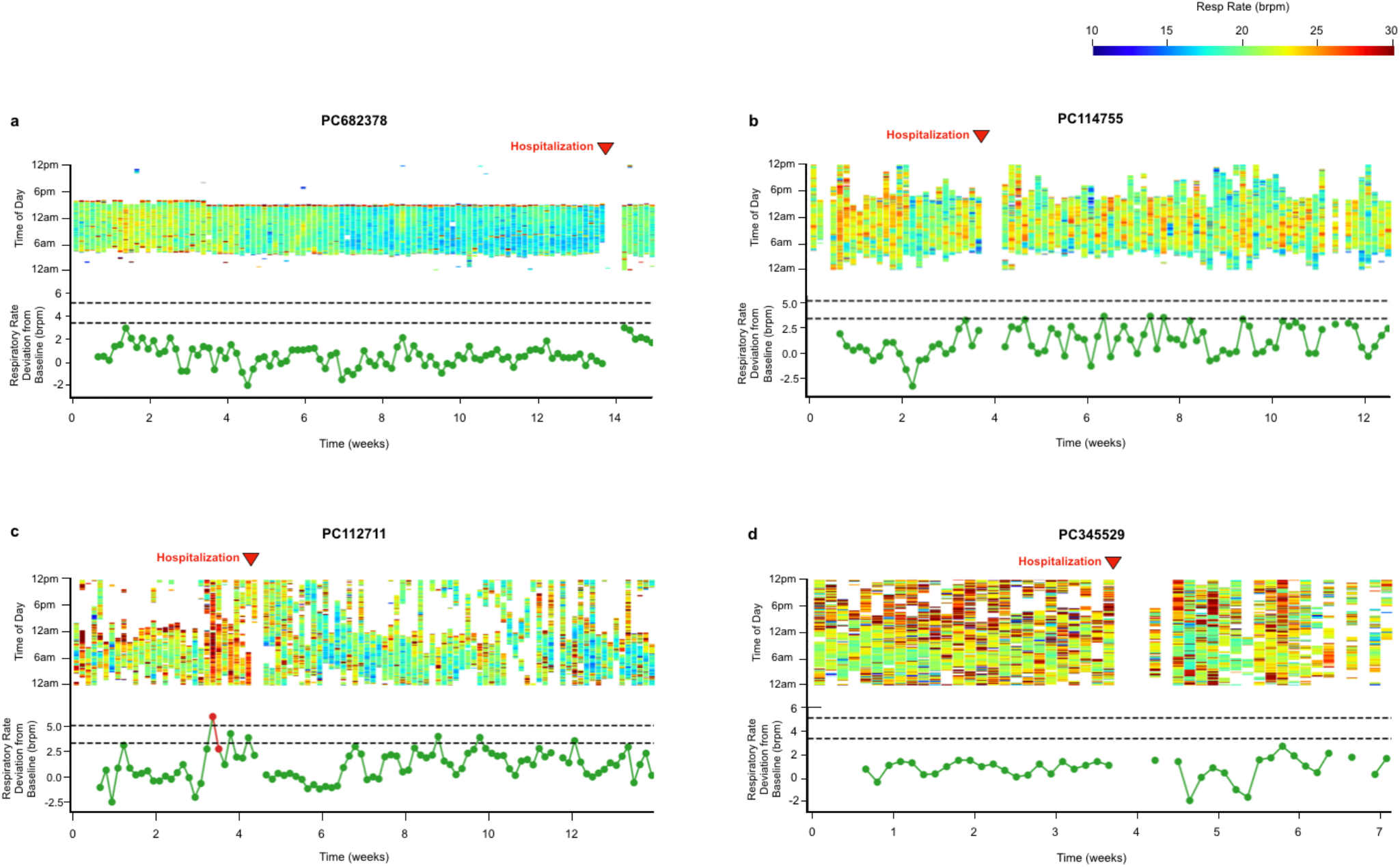
Heatmaps for patients with hospitalization events, NRR not predictive. Heatmap and corresponding NRR deviation from baseline for 4 patients whose hospitalization events were not preceded by a rise in NRR. Heatmap colorbar ranges from 10-30 brpm, values below 10 brpm not shown. NRR excursions colored by low risk (green) and high risk (red). Black dashed lines show RRth (bottom) and RRth+ (top). **a**, Patient was hospitalized for hypoglycemia. **b**, Patient was hospitalized for atrial flutter. **c**, Patient was hospitalized for COVID-19. **d**, Patient was hospitalized for volume overload.

**Supplemental Figure 10.**
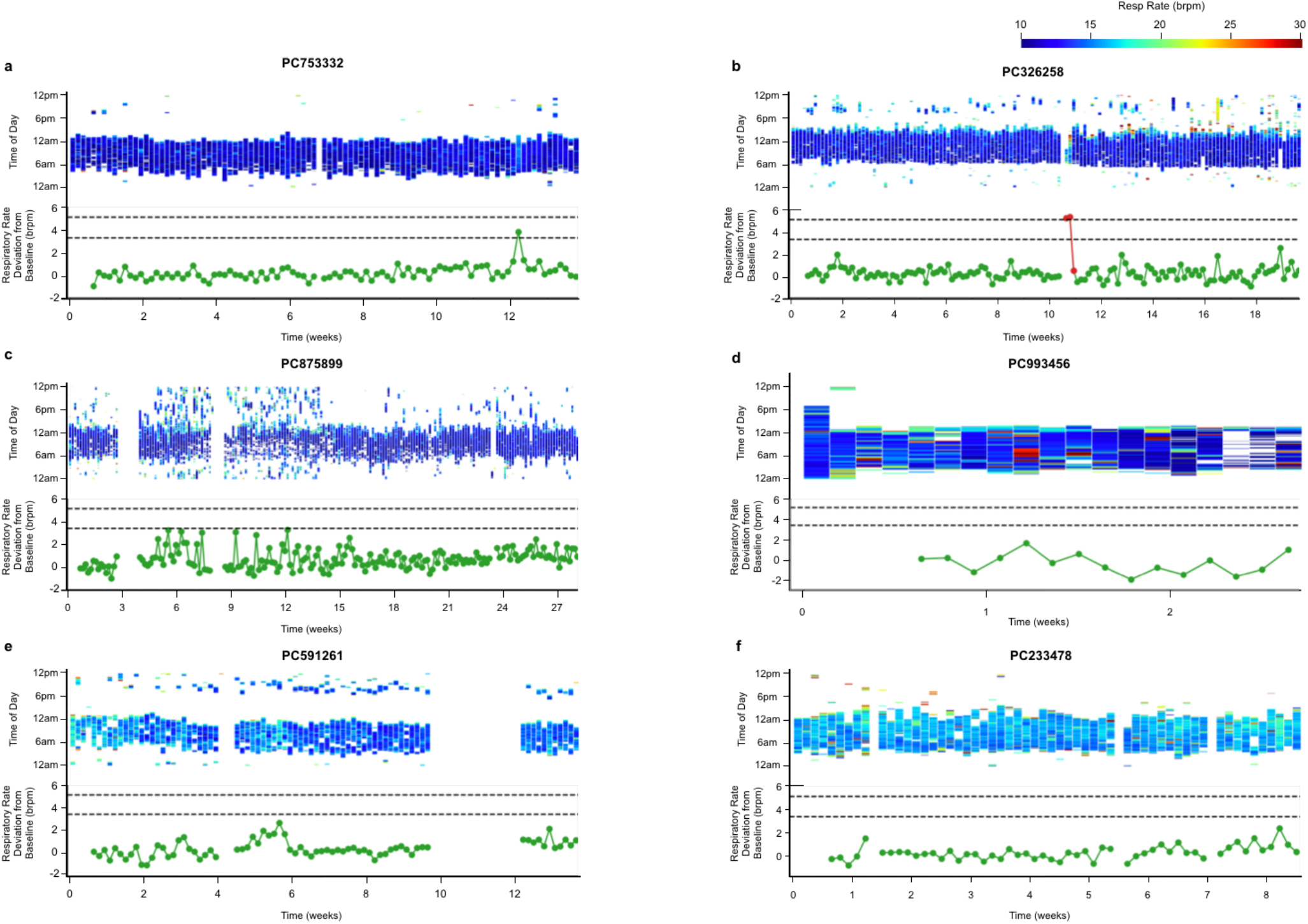
Heatmaps for patients without hospitalization events. Heatmap and corresponding NRR deviation from baseline for 6 patients who did not have hospitalization events during our monitoring period. Heatmap colorbar ranges from 10-30 brpm, values below 10 brpm not shown. NRR excursions colored by low risk (green) and high risk (red). Black dashed lines show RRth (bottom) and RRth+ (top).

**Supplemental Figure 11.**
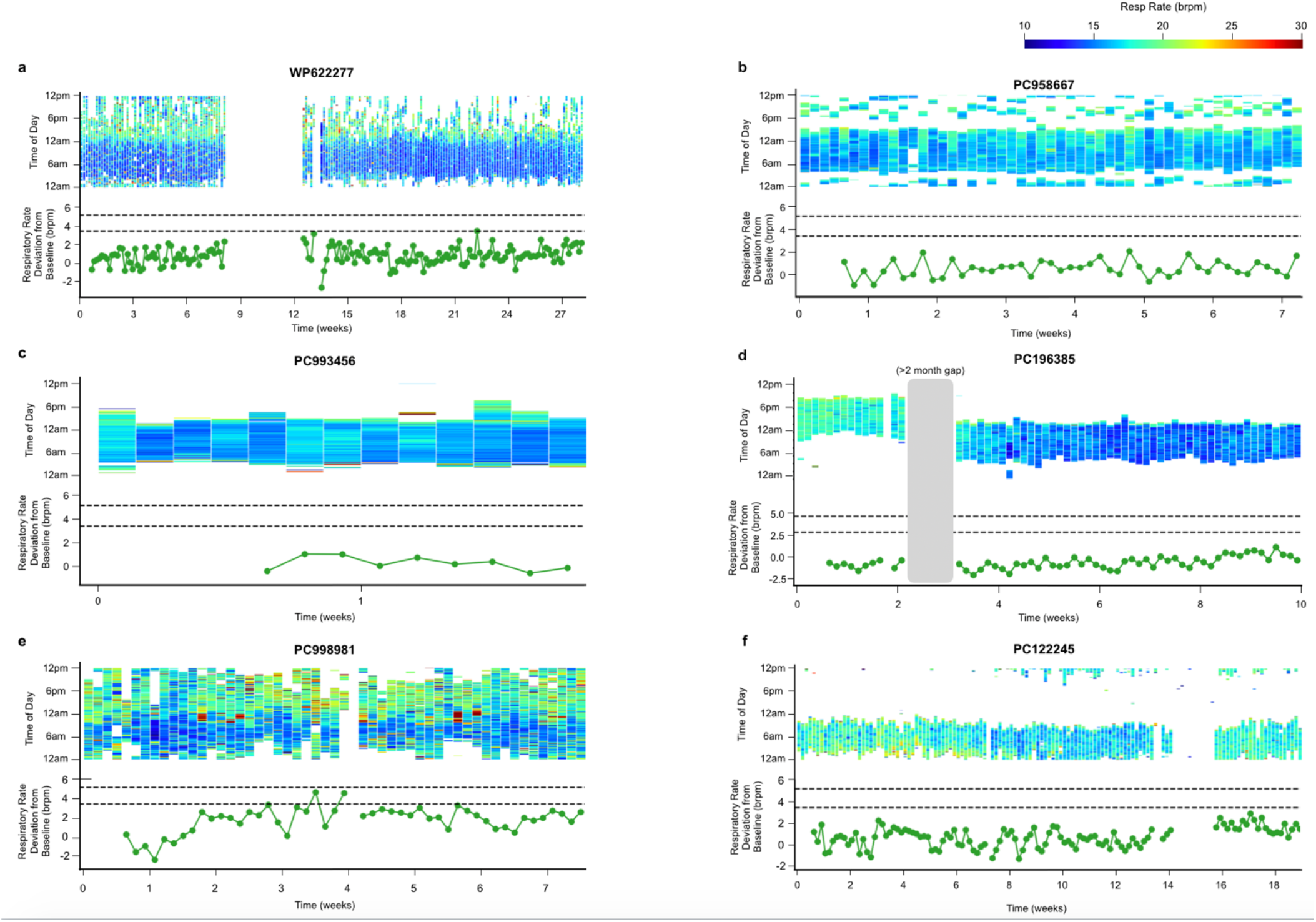
Heatmaps for patients without hospitalization events. Heatmap and corresponding NRR deviation from baseline for 6 patients who did not have hospitalization events during our monitoring period. Heatmap colorbar ranges from 10-30 brpm, values below 10 brpm not shown. NRR excursions colored by low risk (green) and high risk (red). Black dashed lines show RRth (bottom) and RRth+ (top).

**Supplemental Figure 12.**
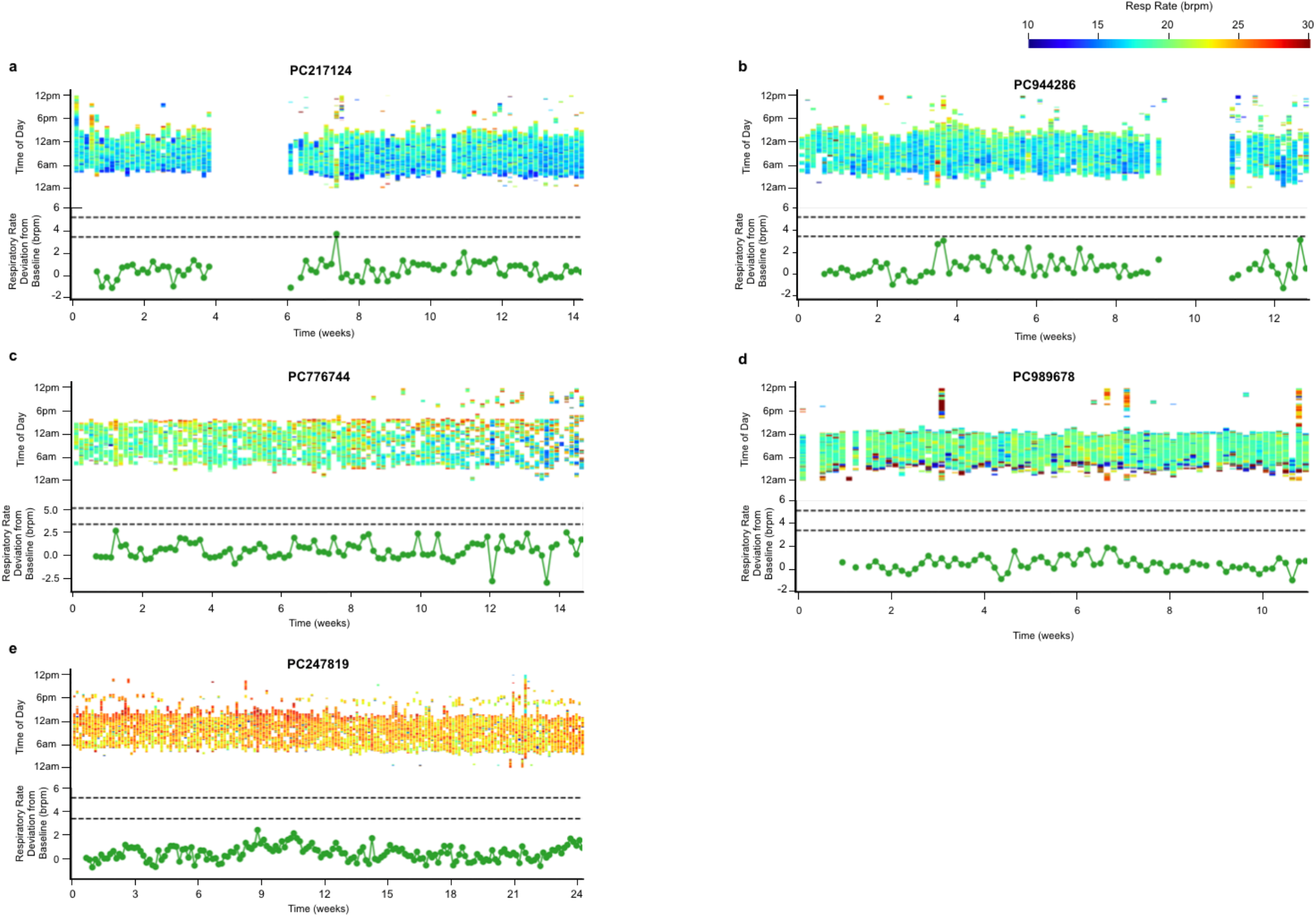
Heatmaps for patients without hospitalization events. Heatmap and corresponding NRR deviation from baseline for 5 patients who did not have hospitalization events during our monitoring period. Heatmap colorbar ranges from 10-30 brpm, values below 10 brpm not shown. NRR excursions colored by low risk (green) and high risk (red). Black dashed lines show RRth (bottom) and RRth+ (top).

